# Geometrically-aggregated training samples: Leveraging summary statistics to enable healthcare data democratization

**DOI:** 10.1101/2023.10.24.23297460

**Authors:** Jenny Yang, Anshul Thakur, Andrew A. S. Soltan, David A. Clifton

## Abstract

Healthcare data is highly sensitive and confidential, with strict regulations and laws to protect patient privacy and security. However, these regulations impede the access of healthcare data to a wider AI research community. As a result, AI healthcare research is often dominated by organisations with access to larger datasets or limited to silo-based development, where models are trained and evaluated on a limited population. Taking inspiration from the non-sensitive nature of the summary statistics (mean, variance, etc.) of healthcare data, this paper proposes *geometrically-aggregated training samples (GATS)* where each training sample is a convex combination of multiple patients’ characteristics. Thus, mappings from patients to any constructed sample are highly convoluted, preserving patient privacy. We demonstrate that these “summary training units” provide effective training on different tabular and time-series datasets (CURIAL, UCI Adult, and eICU), and indeed behave as a summary of the original training datasets. This approach takes important steps towards data accessibility and democratization.

## 1 Introduction

Healthcare data often contains sensitive and confidential information about individuals’ health conditions, medical histories, and demographic information. As such, there are regulations and laws imposing strict requirements on how this data can be collected, used, and shared, requiring that appropriate measures be taken to protect its privacy and security.

These regulations frequently act as barriers to the democratization of healthcare data, making it challenging for artificial intelligence (AI) researchers seeking access to healthcare data. Democratizing healthcare data can bring several benefits, including: 1) creating global clinical models by combining datasets from various locations and diverse populations, enhancing model generalizability and robustness; 2) promoting collaboration between researchers and across healthcare organizations; and 3) enhancing transparency and reproducibility of data-driven algorithms.

Irreversible de-identification or anonymization of electronic health records (EHRs) directly supports data democratization by ensuring that de-identified EHRs cannot be linked to specific patients. However, there is no foolproof de-identification method, and it is always possible to map so-called de-identified EHRs back to individual patients (1; 2; 3).

The objective of this study is to enhance the security of de-identified pre-processed clinical data and facilitate data democratization without compromising the performance of predictive models. We exploit the fact that summary statistics (mean, variance, etc.) commonly shared to describe study cohorts, are not considered sensitive information in healthcare data (3). Since these summary statistics represent population characteristics rather than any individual patient’s features, they are not typically regarded as private or sensitive.

Based on this understanding, we introduce a novel method called “*geometrically-aggregated training samples (GATS)*,” which constructs training samples using convex combinations of multiple patients’ characteristics. Thus, importantly, *GATS* occupies the same data space as real training samples, enabling effective model training. This approach ensures that mappings from patients to any constructed sample become highly intricate, thereby preserving patient privacy. Moreover, the proposed approach can further be augmented with methods such as differential privacy, for further protection against set-membership inference attacks.

To validate the distinctiveness of *GATS* from real training samples, we perform correlation-based quality checks. We evaluate the efficacy of these “summary training units” by applying it to two extensive and data-rich healthcare datasets, namely CURIAL and eICU (which are tabular, and time-series, respectively). Additionally, we assess its performance on a non-healthcare dataset, specifically the UCI Adult dataset, thereby demonstrating its generalizability across diverse domains.

## 2 Related Works

Previous research efforts aimed at addressing data privacy concerns have predominantly focused on three main approaches: creating or utilizing synthetic datasets for model development, differential privacy, and implementing decentralized and distributed model training techniques. However, in this paper, our primary focus is on methods that facilitate data democratization. Consequently, we will not delve into methods specifically related to decentralized and distributed model training, such as federated learning.

Researchers have demonstrated that Generative Adversarial Networks (GANs) can generate large and diverse synthetic data, representative of real-world data(4; 5; 6; 7; 8; 9; 10; 11; 12; 13; 14). Synthetic data generation not only protects patient privacy but also offers a cost-effective means of training AI models, especially when obtaining real-world data is challenging or expensive (e.g., medical images or rare disease cases). Despite the benefits, concerns about data quality arise with synthetic data. Designing, validating, and verifying synthetic data to accurately capture the complexity and variability of real-world data can be a time-consuming and resource-intensive task, requiring expertise and infrastructure. The lack of standardized measures and evaluation metrics for assessing synthetic data quality, particularly in the healthcare domain (with its heterogeneity and variation across populations, diseases, and settings), further complicates the process (15). Additionally, training GANs can be challenging due to potential issues such as overfitting to training data(16; 17), resulting in insufficient diversity in synthetic data; convergence instabilities (18), leading to poor quality synthetic data; and mode collapse (19; 20), which limits the variety of samples produced by the generator, resulting in reduced diversity in the synthetic data.

Differential privacy is an approach where a machine learning (ML) algorithm is designed to safeguard the privacy of individuals whose data is used for training. By integrating differential privacy techniques into the data generation process, synthetic datasets can be created with privacy assurances (10; 11; 13; 21). However, it’s important to note that although differential privacy provides statistical privacy guarantees, the addition of significant noise can reduce the usefulness of the data for ML development, especially when the original data itself may be inherently noisy(22). Moreover, the absence of a standardized approach or metric for implementing differential privacy in ML poses challenges in terms of implementation, evaluation, and practical deployment.

## 3 Proposed Method

Our proposed approach - *GATS* - utilizes summary statistics (such as mean and variance) to generate training samples for data-driven algorithms. Each training unit represents the “summary” of real data points, incorporating the essential information from the original dataset while maintaining the privacy of individual patients. This method addresses the limitations of previous approaches in the following ways:

1. It contains real information (summary statistics), eliminating the need to create synthetic data points (which typically requires substantial amounts of data, resources, and validation efforts).
2. There is no requirement to introduce noise or apply masking/encoding techniques to the data, thereby preserving its usability for learning tasks.
3. It preserves patient privacy, enabling data sharing and democratization by allowing institutions and groups to exchange clinically-rich data.

To construct a new sample (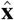, ŷ) for class *C*, we randomly select 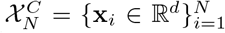 training samples of class *C* and aggregate them using a random convex combination:

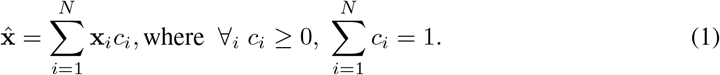

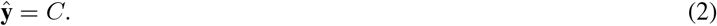

The coefficients *c*_*i*_ are randomly sampled while enforcing the simplical constraint.

Unsurprisingly, we found that class-wise sample generation leads to comparable performances as training on original data at low values of *N* (i.e. when less patient samples are combined together); however, steadily decreases in performance as the value of *N* increases (Section 5, Results). Thus, to both improve class imbalances and regularization during training, we extend the above process to generate mixed-class samples, where *N* training samples from different classes are aggregated to form a new sample. The label of this constructed sample is determined by a majority voting rule on the labels of *N* input examples. Majority voting is also used for any binary/discrete variables.

As such, for each new sample, (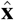, ŷ), 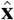 is formed as before (Equation 1); and ŷ is now constructed as:

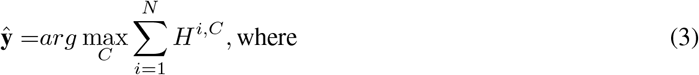

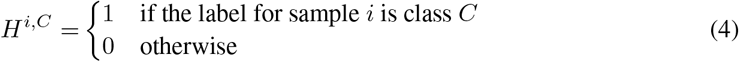

*GATS* replaces the original feature vectors with their interpolated versions which are a function of *N* randomly chosen original feature vectors. Similar to *mixup* augmentation (23) used in computer vision, these interpolated vectors (along with the original examples) expand the training data distribution. However, since we can’t use original examples to avoid patient data leakage, training on *GATS*-generated examples is equivalent to training models on a perturbed version of the original data that encourages implicit regularisation (23; 24; 25; 26), robustness against adversarial attacks (27) and enhances the information bottleneck (28), providing implicit differential privacy to some extent. Moreover, *GATS* can also be used to decrease the class imbalance and prevent overfitting to the training data, as diverse minority class examples can be constructed by aggregating mixed-class examples.

From a geometric standpoint, the *N* chosen training examples can be visualized as the vertices of a simplex. According to the principles of simplex geometry, the constructed sample 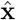 must reside within the convex hull of this simplex. When implemented, it is possible for the constructed sample to be situated near a real training example within the data space. To prevent such situations, we eliminate constructed samples that demonstrate a higher correlation (above 0.75) with real training examples. This additional check guarantees that the publicly released *GATS*-generated data bears no resemblance to any genuine patient samples.

In practical terms, implementing *GATS* is a straightforward process involving three key parameters:

1. The parameter *p* determines the proportion of mixed-label samples to be used.
2. The parameter *q* specifies the mixed-label ratio, which indicates the percentage of samples from class *C*_0_ to mix in when generating a sample from class *C*_1_. For instance, if *N* = 100 and *q* = 0.25, then 25 samples of class *C*_0_ and 75 samples of class *C*_1_ will be utilized to create a single *GATS* sample of class *C*_1_.
3. The parameter *b* defines the batch size employed during the execution of *GATS*. This parameter can be particularly beneficial for managing memory constraints in situations involving extremely large datasets.

By adjusting these three parameters, *GATS* can be easily implemented to suit specific requirements, ensuring efficient execution with minimal computational impact (implementation times can be found in the Supplementary Material).

## 4 Experimental Set-Up

### 4.1 Datasets

We demonstrate our method on two large and clinically-rich datasets (CURIAL, eICU); and additionally, evaluate one non-healthcare dataset (UCI Adult).

#### CURIAL

The CURIAL datasets (29; 30) consist of anonymized electronic health record (EHR) data (including demographic information, blood tests, and vital signs) from emergency departments (EDs) across four independent United Kingdom (UK) National Health Service (NHS) Trusts. These datasets are used for the binary classification task of diagnosing COVID-19.

As CURIAL contains five cohorts across four hospital trusts, we split the PUH cohort into training and internal validation sets, and use the other four cohorts (UHB, BH, OUH “wave 2", and OUH “wave 1”) as external validation sets.

#### UCI Adult (Census Income)

The UCI Adult dataset (31) contains demographic and employment-related features of individuals, such as age, education level, marital status, occupation, and income. It is used for the binary classification task of predicting whether an individual’s income exceeds $50,000 per year.

#### eICU Collaborative Research Database (eICU-CRD)

The eICU dataset (32; 33) is a large, multicenter critical care database comprising of clinical data for ICU admissions from hospitals across the United States. The database includes a variety of data types, such as vital signs, medications, laboratory results, and demographics. In our experiments, this dataset was used for the binary classification task for predicting patient discharge status.

Consistent with previous studies, we addressed any missing values using population median imputation, then standardized features to have a mean of 0 and a standard deviation of 1.

Detailed summary statistics, data availability statements, and ethics approval (where appropriate) can be found in the Supplementary Material.

### 4.2 Metrics

Area under the receiver operator characteristic curve (AUROC) and area under the precision recall curve (AUPRC) are reported, alongside 95% confidence intervals (CIs) based on 1,000 bootstrapped samples taken from the test set. Tests of significance comparing the performance between models are calculated by evaluating how many times one model performs better than another, across 1,000 pairs of bootstrapped iterations.

### 4.3 Baseline and State-of-the-art Comparators

We compare *GATS*-trained neural networks and XGBoost models against equivalent baseline models trained on the corresponding original datasets. This will allow for a 1:1 comparison evaluating the utility of *GATS* for two types of models. We additionally compare our method to current state-of-the-art methods for differentially private, synthetic data generation including marginal (MWEM), neural network (PATE-CTGAN), and hybrid (QUAIL) synthesizers.

#### Multiplicative Weights Exponential Mechanism (MWEM)

The MWEM algorithm utilizes the Multiplicative Weights technique to maintain and update an approximate distribution, through analyzing differences between true and approximate datasets. It uses the Exponential Mechanism (10) to identify the queries that provide the most information to the Multiplicative Weights algorithm. MWEM has been shown produce differentially private synthetic data which match theoretical accuracy guarantees.

#### Conditional Tabular GAN using Private Aggregation of Teacher Ensembles (PATE-CTGAN)

PATE-GAN (11) is a state-of-the-art privacy-preserving machine learning technique that combines two methods: Generative Adversarial Networks (GANs) and Private Aggregation of Teacher Ensembles (PATE). The GAN is used to generate synthetic data, while PATE is used to preserve the privacy of the original data. Additionally, PATE-GAN is capable of generating mixed-type (continuous, discrete, and binary) variables. We combine this with a conditional tabular GAN (CTGAN), which is a GAN-based method specifically designed to improve synthetic data creation for tabular data (12).

#### Quailified Architecture to Improve Labeling (QUAIL)

QUAIL (13) is a method that combines a differentially private classifier and a differentially private synthesizer to generate synthetic data. It fits the classifier on the original data, then uses the synthesizer to learn the distribution of the feature columns from the original data. In our experiments, we use the PATE-CTGAN as the synthesizer backbone. Synthetic data is then generated by sampling feature rows from the fitted synthesizer and generating labels using the previously learned classifier. This approach allows practitioners to control the privacy budget spent on classification versus learning the feature distribution.

It should be noted that MWEM was only tested on the Adult dataset as (10) specifically focused on categorical attributes. Since CURIAL and eICU datasets contain a diverse range of continuous variables, MWEM was not tested on them.

Furthermore, we reference baselines and state-of-the-art results reported in other studies, which utilized the same datasets as presented in this work.

### 4.4 Training Outline

Each model in the study underwent a standardized training and evaluation process. The training set was utilized for various stages, including model development, hyperparameter selection, and model training. A separate validation set was used for continuous validation. Finally, held-out test sets were used to assess the performance of all finalized models.

During model training, the data used consisted of samples generated by the *GATS* method, while the evaluation of models was conducted on the original, real datasets. All baselines and state-of-the-art comparisons are also tested on real data. Results from baseline and state-of-the-art models are implemented and trained ourselves unless otherwise referenced.

## 5 Results

### 5.1 CURIAL Datasets

#### 5.1.1 *GATS* Distributions

By comparing the feature distributions of the original data with the data generated by *GATS*, we can observe that the distribution of features of constructed examples behave like the summary of the distribution of original features, indicating that *GATS* effectively are the statistical summary of the original dataset (see Figure 1). For example, for oxygen saturation, the median and IQR for the original and *GATS*-generated datasets are both 0.142 (−0.110-0.393); for CRP, it’s -0.477 (−0.583-0.109) and -0.437(−0.570-0.189) for the original and *GATS*-generated datasets, respectively; and for haematocrit, it’s 0.086 (−0.551-0.657) and 0.070 (−0.469-0.576) for the original and *GATS*-generated datasets, respectively. Additionally, we notice that the *GATS*-generated samples tend to bring the features closer to their mean, which can be seen as an implicit form of regularization (25). It should be noted that *GATS* was performed on the standardized datasets. Exact values may differ slightly between different implementations of *GATS*, as samples and coefficients are randomly sampled. While the original and *GATS*-generated datasets exhibit similarities in their summary statistics, all features displayed a significant difference in population medians (as determined by the Kruskal-Wallis test, with *p<* 0.05).

**Figure 1:**
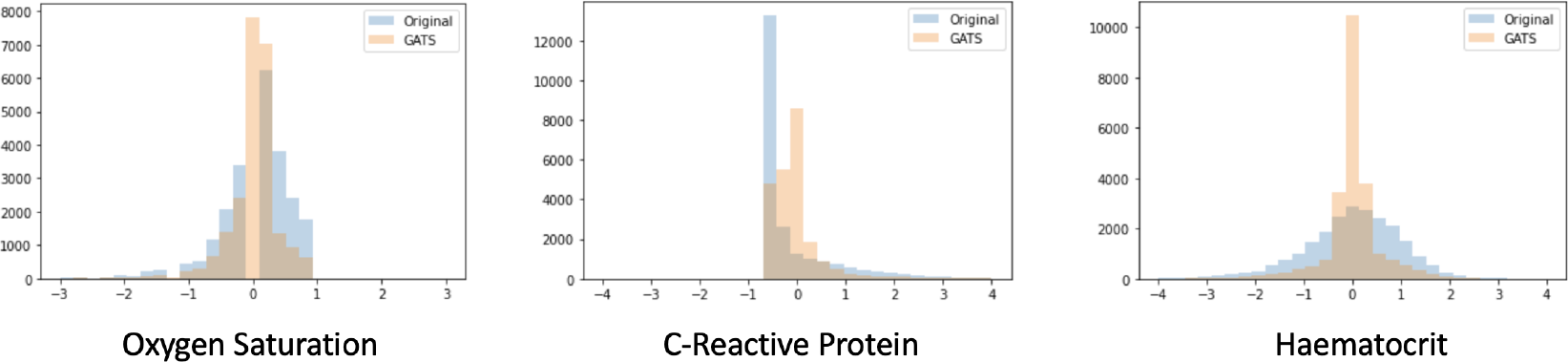
Feature distribution comparison of the original patient dataset and the *GATS*-generated dataset.

In addition, we employed t-Stochastic Neighbor Embedding (t-SNE) to visualize a lower-dimensional representation of both the original data and the *GATS*-generated data for both the COVID-19 positive and negative subgroups (refer to Figure 2). The t-SNE plots reveal the presence of two distinct clusters, indicating that as claimed, the *GATS*-generated data is indeed the perturbed version of the original data.

**Figure 2:**
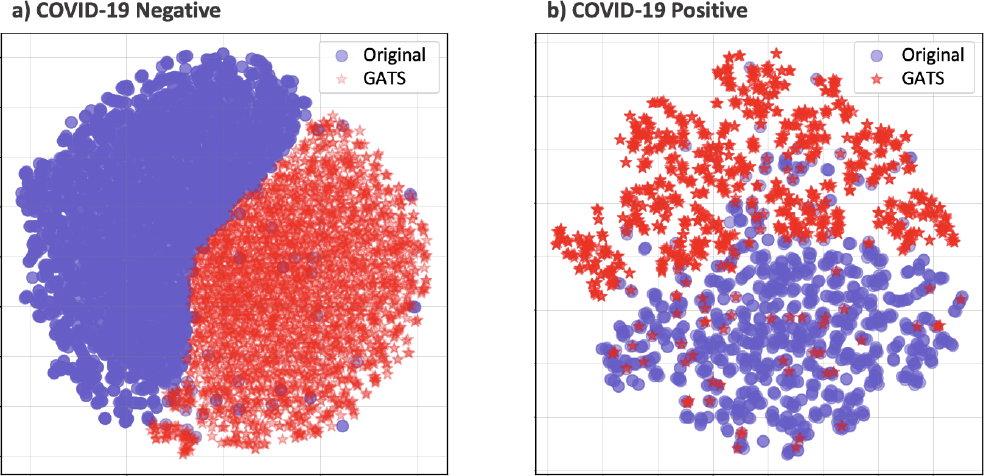
t-SNE comparison of the original patient dataset and the *GATS*-generated dataset, for COVID-19 Negative (a) and Positive (b) samples.

This is further confirmed by the distribution of correlation coefficients comparing original patient samples to *GATS*-generated samples (for *N* =5 and *N* =100), whereby the vast majority of original samples have a correlation magnitude of *<* 0.5 to *GATS* samples (Figure 3). These figures confirm that the privacy of individual patients is preserved through using *GATS*, even at small values of *N*.

**Figure 3:**
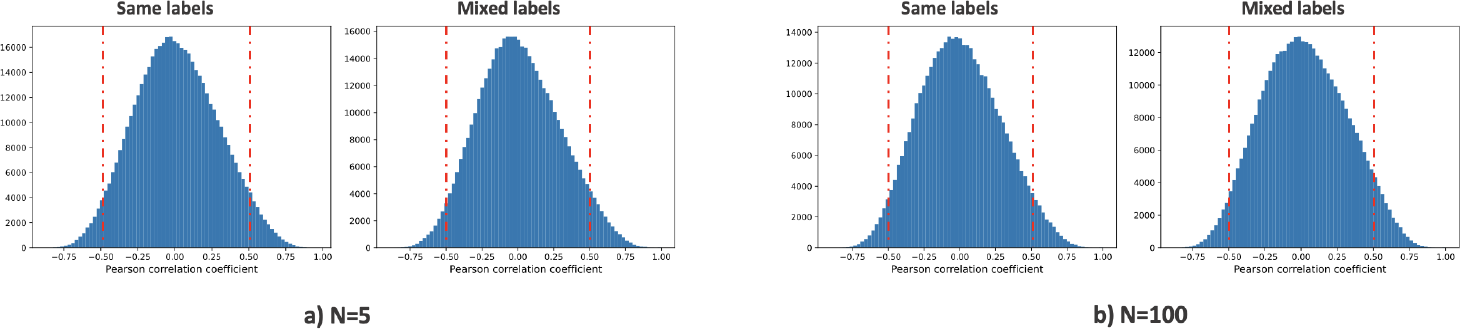
Correlations between same- and mixed-label *GATS*-generated samples for (a) *N* =5, and (b) *N* =100. Red dotted-lines mark magnitudes of 0.5 correlation.

#### 5.1.2 Classification Results

Using the CURIAL datasets, we assessed the effectiveness of training models using class-wise *GATS* compared to mixed-label *GATS* data. Figure 4 (a) demonstrates that when solely utilizing class-wise *GATS* data without any mixed-label samples, the predictive performance (using a neural network) generally decreased as the privacy level (i.e., the number of combined patients, *N*) increased. However, Figure 4 (b) illustrates that incorporating mixed-label *GATS* data (with parameters *p*=0.25 and *q*=0.3) into the training process maintained consistent performance, comparable to a model trained on the original dataset (baseline performance on the original dataset is represented by *N* =1). The difference in accuracy between training on mixed-label *GATS* and same-label *GATS* was found to be statistically significant (*p<* 0.0001) for all values of *N*. This trend was consistent across all five test sets. Similar outcomes were obtained when training an XGBoost model, further validating that the *GATS*-generated data retained the essential information from the original dataset. Comprehensive numerical results for the neural network and XGBoost models can be found in the Supplementary Material.

**Figure 4:**
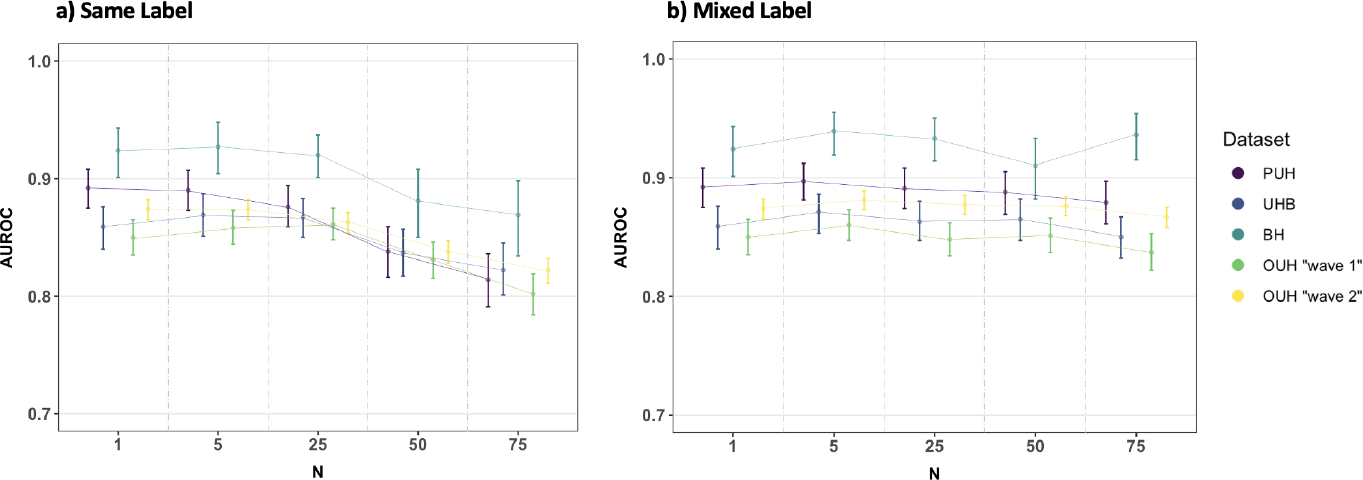
AUROC performances using a neural network-based model, alongside 95% confidence intervals, for different values of N for (a) class-wise *GATS* and (b) mixed-class *GATS* (*p*=0.25, *q*=0.3). Full numerical results for both AUROC and AUPRC can be found in the Supplementary Material.

#### 5.1.3 Ablation Study

We additionally studied the effects of the key hyperparameters for *GATS*, at patient combination levels of *N* =5, *N* =50, and *N* =100. We evaluate the mixed label ratio *q*, the batch size *b*, and the proportion of mixed label *GATS* used in training *p*.

Figure 5 shows the validation AUROC across different privacy levels, *N*. For different mixed label ratios, *q*, we present results for constructing COVID-19 positive samples using a mixture of both COVID-19 positive and negative patients. The COVID-19 positive class was selected due to its status as the minority class within the datasets, which increases the likelihood of overfitting. Generally, the performance remains relatively stable at *N* =5 across all five test sets. However, for larger *N* values, it is observed that utilizing a higher proportion of COVID-19 negative patients in each mixed-label sample enhances performance. Similarly, at higher *N* values, increasing the proportion of mixed-label *GATS* data within the overall training set, *p*, appears to have a slight positive impact on predictive performance. These performance improvements are expected since the mixed-label samples (both within and across individual *GATS*-generated samples), contribute to increased regularization during training. Batch size does not significantly affect the predictive performance of the generated samples; however, this may have impact on much larger datasets (e.g. *»* 10^5^ samples).

**Figure 5:**
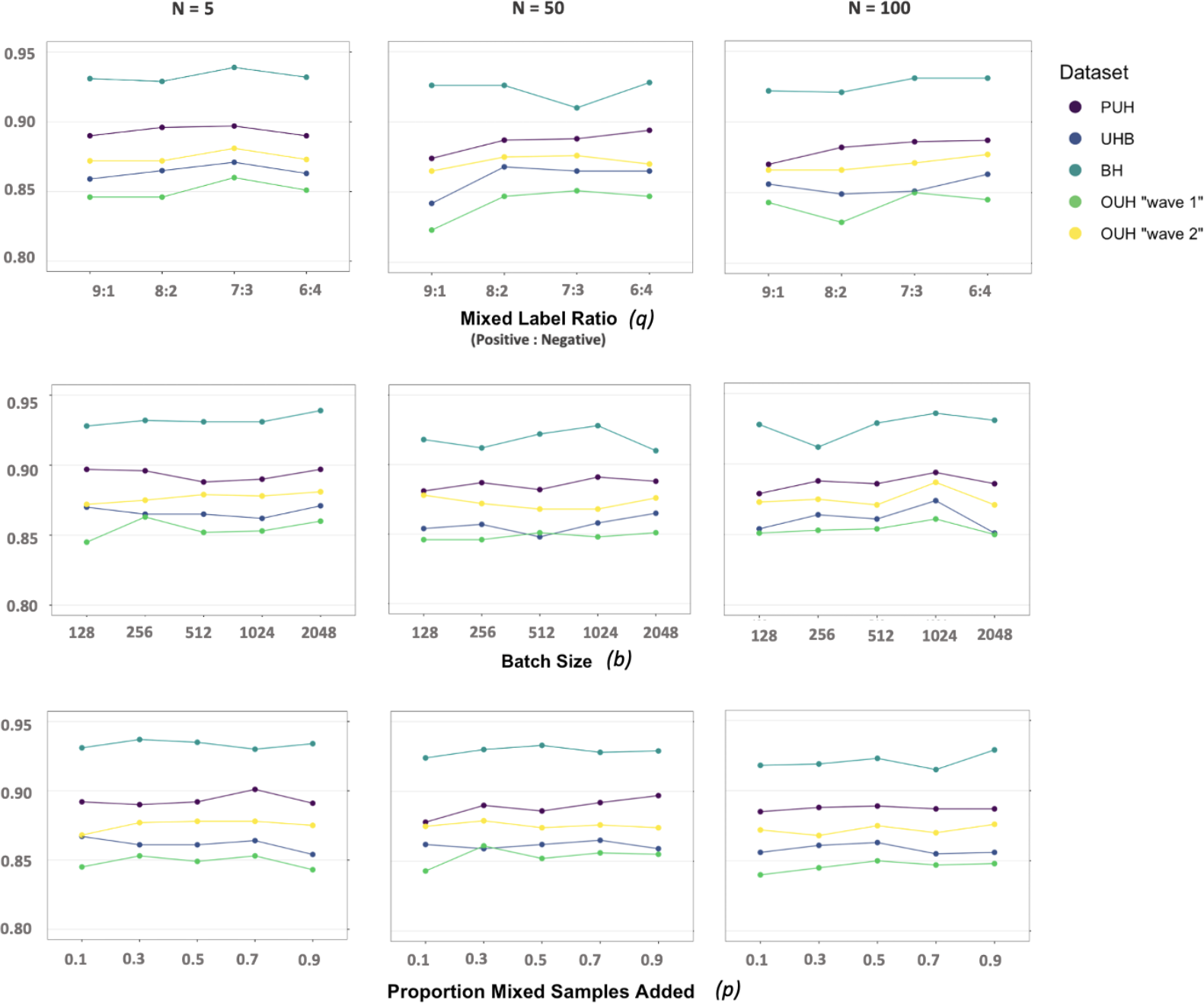
Ablation study of the mixed label ratio *q*, the batch size *b*, and the proportion of mixed label *GATS* added in training *p*. Performance metric shown is AUROC. Full numerical results, alongside 95% confidence intervals can be found in the Supplementary Material.

#### 5.1.4 Baseline and State-of-the-art Comparisons

Table 1 presents results of *GATS* training sets generated using different values of *N*. Results are for *GATS*-trained neural network models, as this was the best performing baseline we trained. Results for corresponding XGBoost models were similar, and can be found in the Supplementary Material.

**Table 1:** Baseline and state-of-the-art comparisons of AUROC performances (alongside 95% CIs, unless otherwise stated), for the CURIAL datasets. Red and blue values denote best and second best scores, respectively. Comparator studies marked with (*) considered data privacy-preservation.

| Method | PUH | UHB | BH | OUH "wave 2" | OUH "wave 1" |
| --- | --- | --- | --- | --- | --- |
| *GATS* (N=5) | **0.897(0.881-0.912)** | **0.871(0.853-0.886)** | **0.939(0.919-0.955)** | **0.881(0.873-0.889)** | **0.860(0.847-0.873)** |
| *GATS* (N=25) | 0.891(0.874-0.908) | 0.863(0.847-0.880) | 0.933(0.914-0.950) | 0.877(0.869-0.885) | 0.848(0.834-0.862) |
| *GATS* (N=50) | 0.888(0.869-0.905) | 0.865(0.847-0.882) | 0.910(0.882-0.933) | 0.876(0.868-0.884) | **0.851(0.837-0.866)** |
| *GATS* (N=75) | 0.879(0.861-0.897) | 0.850(0.832-0.867) | **0.936(0.915-0.954)** | 0.867(0.858-0.875) | 0.837(0.822-0.853) |
| State-of-the-art comparisons (synthetic data) |  |  |  |  |  |
| QUAIL | 0.823(0.801-0.845) | 0.821(0.802-0.839) | 0.893(0.868-0.916) | 0.812(0.802-0.820) | 0.789(0.772-0.806) |
| PATE-CTGAN | 0.573(0.550-0.599) | 0.583(0.560-0.605) | 0.482(0.443-0.524) | 0.607(0.596-0.618) | 0.607(0.589-0.625) |
| Baseline comparisons (original data) |  |  |  |  |  |
| Neural Network | **0.892(0.875-0.908)** | 0.859(0.840-0.876) | 0.924(0.901-0.943) | 0.874(0.865-0.882) | 0.850(0.835-0.865) |
| XGBoost | **0.892(0.873-0.908)** | 0.841(0.823-0.859) | 0.914(0.890-0.933) | 0.856(0.847-0.865) | 0.818(0.802-0.834) |
| XGBoost (30) | 0.872(0.863-0.882) | 0.858(0.838-0.878) | 0.881(0.851-0.912) | **0.878(SD 0.001)** |  |
| *Neural Network (34) | 0.857 | 0.864 | 0.861 |  |  |
| Neural Network (35) | 0.867(0.857-0.877) | **0.867(0.845-0.888)** | 0.894(0.859-0.929) | 0.866(0.855-0.876) |  |
| Reinforcement Learning (36) | 0.831(0.819-0.842) | 0.837(0.814-0.861) | 0.867(0.829-0.906) | 0.861(0.850-0.871) |  |
| *Federated Learning (37) | 0.876(0.865-0.886) |  | 0.917(0.893-0.942) | 0.872(0.862-0.882) |  |

As presented in Table 1, *GATS* with *N* =5 consistently outperformed state-of-the-art and baseline comparators in terms of AUROC performance across all test sets. Even when using *GATS* generated at higher values of *N*, the models still achieve comparable performances to baselines trained on the original data. For instance, when *N* =75, the maximum decrease in AUROC is only 0.013 compared to the corresponding neural network baseline. In contrast, the classification results obtained from synthetic data generated by state-of-the-art QUAIL and PATE-CTGAN models did not surpass the performance of *GATS* across all test sets and values of *N*.

### 5.2 eICU-CRD

In the case of the eICU dataset, we observed a general decrease in predictive performance as the privacy parameter *N* increased when using only class-wise *GATS*. However, similar to our previous findings, the introduction of mixed-label *GATS* led to consistent and comparable performance to the baseline model trained on the original dataset. This pattern was observed for both neural network and XGBoost models (complete numerical results can be found in the Supplementary Material). The difference in accuracy between training on mixed-label *GATS* and same-label *GATS* was statistically significant (p*<*0.0001) for all values of *N*.

Table 2 presents the results of training models on *GATS* data, generated with different values of *N*, using *p*=0.25 and *q*=0.3. The reported results are for XGBoost models, which performed the best among the baseline models. Similar results were obtained for the corresponding neural network models, and they can be found in the Supplementary Material.

**Table 2:** Baseline and state-of-the-art comparisons of AUROC and AUPRC performances (alongside 95% CIs, unless otherwise stated), for the eICU dataset. Red and blue values denote best and second best scores, respectively.

| Method | AUROC | AUPRC |
| --- | --- | --- |
| *GATS* (N=5) | 0.864(0.857-0.872) | 0.495(0.475-0.517) |
| *GATS* (N=25) | 0.864(0.857-0.872) | 0.494(0.474-0.516) |
| *GATS* (N=50) | 0.864(0.856-0.872) | 0.499(0.478-0.52) |
| *GATS* (N=75) | 0.864(0.857-0.872) | 0.496(0.475-0.519) |
| *GATS* (N=100) | **0.867(0.860-0.875)** | 0.501(0.482-0.522) |
| State-of-the-art comparisons (synthetic data) |  |  |
| QUAIL | 0.715(0.704-0.727) | 0.207(0.195-0.220) |
| PATE-CTGAN | 0.595(0.584-0.608) | 0.154(0.144-0.167) |
| Baseline comparisons (original data) |  |  |
| Neural Network | 0.851(0.843-0.860) | 0.473(0.453-0.495) |
| XGBoost | **0.879(0.872-0.886)** | **0.527(0.507-0.549)** |
| Logistic Regression (LASSO) (38) | 0.820 | 0.208 |
| BiLSTM (39) | 0.866 (0.66) | **0.552** |

Across different values of *N*, the models trained on *GATS* performed consistently. Although slightly lower, the AUROC values were only 0.012-0.015 lower compared to the XGBoost baseline, indicating that the *GATS* data captures the essential information from the original dataset.

When comparing results to synthetic data generated by state-of-the-art QUAIL and PATE-CTGAN models, the performance of *GATS* consistently outperformed them across all values of *N*.

### 5.3 Adult (Census Income)

When applying *GATS* to the Adult dataset, we observed a decrease in predictive performance as privacy, *N*, increased when using class-wise *GATS* exclusively. However, once mixed-label *GATS* samples were incorporated, the classifier achieved consistent performance, comparable to a baseline model trained on the original dataset. This pattern was observed for both neural network and XGBoost models, and detailed numerical results can be found in the Supplementary Material. The difference in accuracy between training on mixed-label *GATS* and same-label *GATS* was statistically significant (p*<*0.0001) for all values of *N*.

Table 3 presents *GATS*-based (results presented use *p*=0.4, and *q*=0.3) results for XGBoost models, as this was the best performing baseline achieved. Results for *GATS*-trained neural network models were similar, and can be found in the Supplementary Material. Using *GATS*-generated data, models appear to perform consistently across varying values of *N*. AUROC scores were only 0.015-0.027 lower for all *GATS*-trained models compared to the XGBoost baseline, confirming that *GATS* data retains the underlying information from the original dataset.

**Table 3:** Baseline and state-of-the-art comparisons of AUROC and AUPRC performances (alongside 95% CIs, unless otherwise stated), for the Adult dataset. Red and blue values denote best and second best scores, respectively.

| Method | AUROC | AUPRC |
| --- | --- | --- |
| *GATS* (N=5) | 0.909(0.904-0.915) | 0.789(0.774-0.802) |
| *GATS* (N=25) | 0.911(0.905-0.917) | 0.790(0.776-0.804) |
| *GATS* (N=50) | **0.913(0.907-0.919)** | 0.797(0.784-0.811) |
| *GATS* (N=75) | **0.913(0.907-0.919)** | 0.796(0.783-0.810) |
| *GATS* (N=100) | **0.913(0.908-0.919)** | **0.798(0.785-0.811)** |
| State-of-the-art comparisons (synthetic data) |  |  |
| QUAIL | 0.708(0.697-0.720) | 0.448(0.429-0.468) |
| MWEM | 0.599(0.586-0.610) | 0.286(0.274-0.299) |
| PATE-CTGAN | 0.506(0.503-0.509) | 0.236(0.227-0.244) |
| IT-GAN (14) | 0.86 |  |
| CTGAN (12) | 0.85 (Accuracy) |  |
| Baseline comparisons (original data) |  |  |
| Neural Network | 0.888(0.882-0.895) | 0.736(0.722-0.753) |
| XGBoost | **0.928(0.922-0.933)** | **0.826(0.814-0.839)** |

Furthermore, our findings showed that *GATS* outperformed QUAIL, MWEM, and PATE-CTGAN, as well as other GAN models trained on the same dataset, across all values of *N*.

## 6 Conclusion and Discussion

In this paper, we introduce a novel framework called *GATS* that generates informative training samples by combining features from multiple patients. Thus, the fabricated samples exist within the same data space as the real training samples, enabling effective model training. These generated samples can be seen as a “summary” of multiple patients (as opposed to “synthetic” data) and may not be subject to the same level of sensitivity as individual patient data, potentially alleviating concerns related to data privacy regulations. The difficulty in mapping a generated sample to a specific patient enables data sharing and represents an initial step towards healthcare data democratization.

One important consideration is that the performance of *GATS* is influenced by its hyperparameters, as indicated by our findings. The results clearly demonstrate that the choice of *N, p*, and *q* significantly impacts performance. Therefore, future research can explore practical methods for setting these hyperparameters without the necessity of conducting an exhaustive hyperparameter sweep or grid search, which can be computationally demanding. Particularly in environments such as hospitals where computational resources may be limited, the *GATS* method may add complexity to the data processing stage. Some viable alternatives include employing random search, which entails randomly selecting hyperparameters from predefined ranges. This approach is more efficient than a grid search as it explores a wider range while requiring fewer iterations. Another option is to leverage libraries that offer automated tools for hyperparameter optimization. These libraries often incorporate sophisticated techniques like Bayesian optimization or evolutionary algorithms. Nonetheless, it’s essential to acknowledge that determining the optimal hyperparameters may vary based on the specific problem, the particular dataset being used, and the algorithm under consideration.

Similarly, it would be valuable to gain insights into the scenarios where *GATS* demonstrate optimal performance and where it encounters limitations, extending our analysis beyond hyperparameters. Considering real datasets, which often exhibit noise, outliers, and missing data that necessitate imputation, it will be important to investigate how would these issues affect the performance of *GATS*. For example, could outliers significantly disrupt the convex combination? It would be beneficial to assess such considerations from a practical, real-world usage perspective. Particularly in the domain of medical data, there exists a spectrum of case severities. Critical cases, such as those involving patients with various comorbidities, can introduce noticeable fluctuations in data values and deviations from the mean or median. While extreme values can indeed provide valuable insights, it is essential to manage them appropriately to prevent any adverse impacts on model performance. In our conducted experiments, we adhered to the same data preprocessing protocol as employed in prior studies, which did not involve any extreme or improbable outliers. Therefore, for future research, it may prove advantageous to explore additional filtering and preprocessing steps aimed at identifying anomalies and enhancing the dataset’s quality prior to commencing model development and testing.

Another limitation of this study lies in the depth of fidelity assessment. Synthetic data evaluation typically encompasses both utility (pertaining to downstream performance) and fidelity (pertaining to resemblance to real data). In this paper, the primary emphasis is placed on utility and practical application within experiments. However, in contexts involving medical data, where concerns regarding interpretability are likely, fidelity assumes paramount significance. Through the process of summarizing the original data into statistics like mean and variance, the proposed approach (*GATS*) may fail to capture the intricate details and subtleties inherent in the original dataset. While summary statistics offer a high-level overview of the data, they may fall short in encapsulating the full spectrum of diversity and complexity present in individual patient information. Additionally, since the approach generates samples that are combinations, there is potentially a loss of interpretability. For instance, it can become challenging to ensure that the generated samples incorporate feature combinations that align with clinical plausibility. Although the concept of using “summary statistics” maintains the distribution of the real data, it will still be important to further validate the validity and realism of the samples generated. Nevertheless, in situations where the goal is to train models or comprehend the data distributions used in model training, sharing *GATS*-generated data may suffice. This can enable external researchers to replicate studies or gain insights into data disparities during domain adaptation.

To evaluate the distinction between *GATS*-generated samples and the original data in the context of privacy analysis, we employed empirical techniques such as correlation analysis and t-SNE analysis. However, it is imperative to conduct additional experiments in order to develop a more comprehensive understanding of the potential privacy risks and vulnerabilities associated with the proposed approach. For instance, it would be beneficial to explore tasks such as reverse-engineering *GATS*-generated samples to ascertain individual patients’ data or subjecting *GATS* to adversarial attacks. These specific types of attacks fall outside the scope of our current investigations, as our primary focus was directed towards ensuring that samples generated based on “summary statistics” could not be traced back to individual patient samples.

Furthermore, numerous research initiatives have directed their efforts towards mitigating data privacy concerns through decentralized and distributed model training approaches. In our study, we concentrated primarily on promoting data democratization, allowing data to be shared, a concept that decentralized training aims to circumvent. Nevertheless, techniques like federated learning, which involve decentralized and distributed model training, have exhibited impressive performance while enhancing privacy assurances. Consequently, future experiments could explore the integration of various privacy-preserving methods, including differential privacy, *GATS*, and federated learning, to reinforce privacy and facilitate collaborative approaches in the development of machine learning tools.

In this study, we delve into and illustrate the practicality of *GATS* within the medical domain, specifically focusing on its application to electronic health records, which are organized in a tabular data format. Furthermore, we showcase its efficacy using the Adult Census dataset, which falls outside the realm of the medical domain but is also structured as tabular data. However, it’s important to acknowledge that both healthcare and non-healthcare domains encompass a variety of data structures, including images, text, and time-series data. In these alternative scenarios, the application of *GATS* may present challenges due to the potential complexity in interpreting the combined samples produced. For instance, determining how to meaningfully merge doctor’s written notes through a convex combination or how to combine multiple images in a manner that maintains medical plausibility can prove to be non-trivial tasks. Thus, although *GATS* can be applied to different fields, more consideration needs to be given to the data modality being considered. Furthermore, it could be of interest to assess the performance of *GATS* on unlabeled datasets, particularly in the context of unsupervised learning tasks, an area we have yet to explore, having demonstrated *GATS* on supervised learning tasks.

Despite the innovative approach to safeguarding privacy, there may still exist regulatory concerns and obstacles to surmount, given the stringent laws and regulations governing healthcare data. If mishandled, the convolution of patient data could inadvertently lead to unintended consequences, such as the creation of training samples that do not accurately represent specific patient demographics. Moreover, it’s worth emphasizing that while this paper provides a technical solution to address certain legal aspects, it remains essential to involve legal experts and regulatory authorities for a comprehensive assessment of the proposed approach. Such an analysis would serve to uncover any potential legal issues and ensure alignment with prevailing data privacy laws, thereby confirming that the samples generated by *GATS* do not qualify as sensitive information within the current legal framework.

## Data Availability

Data from UHB, PUH and BH are available on reasonable request to the respective trusts, subject to HRA requirements. Data from OUH studied here are available from the Infections in Oxfordshire Research Database (https://oxfordbrc.nihr.ac.uk/research-themes/modernising-medical-microbiology-and-big-infection-diagnostics/infections-in-oxfordshire-research-database-iord/), subject to an application meeting the ethical and governance requirements of the database. Data from UHB, PUH and BH are available on reasonable request from the respective trusts, subject to HRA requirements.
The eICU Collaborative Research Database is available online at https://www.physionet.org/content/eicu-crd/2.0/.
The UCI Adult (Census Income) dataset is available online at https://archive.ics.uci.edu/dataset/2/adult.

https://www.physionet.org/content/eicu-crd/2.0/

https://archive.ics.uci.edu/dataset/2/adult

## Contributions

JY conceived and ran the experiments. JY and AT wrote and implemented the code. JY and AAS preprocessed the COVID-19 datasets. JY preprocessed the eICU and Adult datasets. All authors revised the manuscript.

## Acknowledgements

We express our sincere thanks to all patients and staff across the four participating NHS trusts; Oxford University Hospitals NHS Foundation Trust, University Hospitals Birmingham NHS Trust, Bedfordshire Hospitals NHS Foundations Trust, and Portsmouth Hospitals University NHS Trust.

## Ethics

United Kingdom National Health Service (NHS) approval via the national oversight/regulatory body, the Health Research Authority (HRA), has been granted for use of routinely collected clinical data to develop and validate artificial intelligence models to detect Covid-19 (CURIAL; NHS HRA IRAS ID: 281832).

## Data Availability

Data from UHB, PUH and BH are available on reasonable request to the respective trusts, subject to HRA requirements. Data from OUH studied here are available from the Infections in Oxfordshire Research Database (https://oxfordbrc.nihr.ac.uk/research-themes/modernising-medical-microbiology-and-big-infection-diagnostics/infections-in-oxfordshire-research-database-iord/), subject to an application meeting the ethical and governance requirements of the database. Data from UHB, PUH and BH are available on reasonable request from the respective trusts, subject to HRA requirements.

The eICU Collaborative Research Database is available online at https://www.physionet.org/content/eicu-crd/2.0/.

The UCI Adult (Census Income) dataset is available online at https://archive.ics.uci.edu/dataset/2/adult.

## Code Availability

Code for this will be available upon publication.

