## Supplementary Material for "Geometrically-aggregated training samples: Leveraging summary statistics to enable healthcare data democratization"

#### A Software Packages and Implementation

Models were implemented using Python (v3.6.9) and PyTorch (v1.10.1+cu102). All models were run using an Intel Xeon E-2146G Processor (CPU: 6 cores, 4.50 GHz max frequency).

QUAIL, MWEM, and PATE-CTGAN models were trained using Diffprivlab and Smartnoise packages.

Implementation times:

1. CURIAL (22,737 samples, 26 features): 1.74 s
2. eICU (65,741 samples, 402 features): 47.6 s
3. Adult (24,420 samples, 12 features): 4.17 s

#### B Datasets

##### B.1 CURIAL Datasets

The CURIAL database is an anonymized database with United Kingdom National Health Service (NHS) approval via the national oversight/regulatory body, the Health Research Authority (HRA)(CURIAL; NHS HRA IRAS ID: 281832).

Data from OUH studied here are available from the Infections in Oxfordshire Research Database (https://oxfordbrc.nihr.ac.uk/research-themes/modernising-medical-microbiology-and-big-infection-diagnostics/infections-in-oxfordshire-research-database-iord/), subject to an application meeting the ethical and governance requirements of the Database. Data from UHB, PUH and BH are available on reasonable request to the respective trusts, subject to HRA requirements.

Supplementary Table 1: Clinical predictors considered for COVID-19 status prediction.

| Category | Features |
|---|---|
| Vital Signs | Heart rate, respiratory rate, oxygen saturation, systolic blood pressure, diastolic blood pressure, temperature |
| Blood Tests | Haemoglobin, haematocrit, mean cell volume, white cell count, neutrophil count, lymphocyte count, monocyte count, eosinophil count, basophil count, platelets |
| Liver Function Tests & C-reactive protein | Albumin, alkaline phosphatase, alanine aminotransferase, bilirubin, C-reactive protein |
| Urea & Electrolytes | Sodium, potassium, creatinine, urea, estimated glomerular filtration rate |

Supplementary Table 2: Summary of number of patients, COVID-19 positive cases

| | Training | Validation | Test |
|---|---|---|---|
| Total patients | 22,737 (1,182 positive) | 7,579 (439 positive) | 134,578 (3,680 positive) |
| PUH | 22,737 (1,182 positive) | 7,579 (439 positive) | 7,580 (384 positive) |
| UHB | NA | NA | 10,293 (439 positive) |
| BH | NA | NA | 1,177 (144 positive) |
| OUH "wave 2" | NA | NA | 22,857 (2,012 positive) |
| OUH "wave 1" | NA | NA | 92,671 (701 positive) |

##### B.2 ICU Dataset

The eICU Collaborative Research Database (eICU-CRD) is a publicly-available, anonymized database with pre-existing institutional review board (IRB) approval. The database is released under the Health Insurance Portability and Accountability Act (HIPAA) safe harbor provision. The re-identification risk was certified as meeting safe harbor standards by Privacert (Cambridge, MA) (HIPAA Certification no. 1031219-2).

The eICU Collaborative Research Database is available online at `https://www.physionet.org/content/eicu-crd/2.0/`.

Using similar inclusion and exclusion criteria to those used in previous studies, we selected adult patients (age > 18) with a minimum of 15 ICU records, and grouped these records into 1 hour windows. We removed any samples that did not have a clear discharge status (i.e., anything that was not "alive" or "expired").

Further preprocessing was performed to remove samples with any missing values, one-hot encode categorical features, and standardize all continuous features to have a mean of 0 and a standard deviation of 1.

We used a 80:20 training and test ratio, resulting in 65,741 training (5,987 deaths) and 16,436 (1,486 deaths) test samples, respectively.

Supplementary Table 3: Clinical predictors considered for predicting patient discharge status and patient diagnosis.

| Category | Features |
|---|---|
| Demographic features | Gender, age, height, weight |
| Measurements at hospital admission | Non-invasive systolic blood pressure, non-invasive diastolic blood pressure, non-invasive mean arterial pressure, heart rate, supporting oxygen used at admission, blood oxygen saturation, Glasgow coma score, diagnosis at admission |
| Measurements at ICU admission | Glucose |

##### B.3 Adult (Census Income) Dataset

The UCI Adult (Census Income) Dataset is available online at `https://archive.ics.uci.edu/ml/datasets/Adult/`

#### C Model Architectures

**Neural Network Models**: The rectified linear unit (ReLU) activation function was used for the hidden layers and the Sigmoid activation function was used in the output layer. For updating model weights, binary cross entropy loss and the Adaptive Moment Estimation (Adam) optimizer was used during training. Models were trained using 100 epochs and a batch size of 2048.

Supplementary Table 4: Neural Network hyperparameter settings for COVID-19, income, and discharge status prediction.

| Task | Hyperparameters |
|---|---|
| COVID-19 Diagnosis | n hidden layers = 4<br>n hidden nodes (per layer) = 10<br>learning rate = 0.005 |
| Income Prediction | n hidden layers = 3<br>n hidden nodes (per layer) = 5<br>learning rate = 0.005 |
| Discharge Status Prediction | n hidden layers = 1<br>n hidden nodes (per layer) = 200<br>learning rate = 0.001 |

### D Results

#### D.1 CURIAL datasets

Supplementary Table 5: Comparison of AUROC and AUPRC performances using a neural network-based model (alongside 95% CIs), between same label and mixed label patient sample combinations, for the CURIAL datasets.

| N | Test Set | AUROC | | AUPRC | |
|---|---|---|---|---|---|
| | | Same Labels | Mixed Labels | Same Labels | Mixed Labels |
| Baseline (N=1) | PUH | 0.892(0.875-0.908) | | 0.578(0.536-0.623) | |
| | UHB | 0.859(0.84-0.876) | | 0.359(0.322-0.404) | |
| | BH | 0.924(0.901-0.943) | | 0.723(0.661-0.781) | |
| | OUH "wave 2" | 0.874(0.865-0.882) | | 0.613(0.593-0.633) | |
| | OUH "wave 1" | 0.85(0.835-0.865) | | 0.13(0.112-0.153) | |
| 5 | PUH | 0.890(0.873-0.907) | **0.897(0.881-0.912)** | 0.534(0.489-0.579) | **0.580(0.537-0.623)** |
| | UHB | **0.869(0.851-0.887)** | 0.871(0.853-0.886) | **0.405(0.364-0.455)** | 0.365(0.326-0.405) |
| | BH | 0.927(0.904-0.948) | **0.939(0.919-0.955)** | 0.716(0.648-0.780) | **0.724(0.654-0.790)** |
| | OUH "wave 2" | 0.874(0.865-0.882) | **0.881(0.873-0.889)** | 0.606(0.586-0.627) | **0.633(0.613-0.652)** |
| | OUH "wave 1" | 0.858(0.844-0.873) | **0.860(0.847-0.873)** | **0.139(0.122-0.161)** | 0.129(0.112-0.148) |
| 25 | PUH | 0.876(0.859-0.894) | **0.891(0.874-0.908)** | 0.387(0.347-0.427) | **0.542(0.500-0.591)** |
| | UHB | **0.867(0.850-0.883)** | 0.863(0.847-0.880) | 0.290(0.262-0.322) | **0.317(0.281-0.358)** |
| | BH | 0.920(0.901-0.937) | **0.933(0.914-0.950)** | 0.607(0.536-0.676) | **0.709(0.644-0.777)** |
| | OUH "wave 2" | 0.863(0.854-0.871) | **0.877(0.869-0.885)** | 0.478(0.459-0.496) | **0.588(0.567-0.607)** |
| | OUH "wave 1" | **0.861(0.848-0.875)** | 0.848(0.834-0.862) | **0.082(0.073-0.091)** | 0.072(0.063-0.084) |
| 50 | PUH | 0.838(0.816-0.859) | **0.888(0.869-0.905)** | 0.325(0.290-0.363) | **0.568(0.525-0.615)** |
| | UHB | 0.837(0.817-0.857) | **0.865(0.847-0.882)** | 0.262(0.235-0.290) | **0.343(0.305-0.386)** |
| | BH | 0.881(0.850-0.908) | **0.910(0.882-0.933)** | 0.553(0.487-0.621) | **0.715(0.653-0.769)** |
| | OUH "wave 2" | 0.838(0.828-0.847) | **0.876(0.868-0.884)** | 0.436(0.417-0.453) | **0.630(0.611-0.649)** |
| | OUH "wave 1" | 0.831(0.815-0.846) | **0.851(0.837-0.866)** | 0.065(0.058-0.072) | **0.117(0.101-0.135)** |
| 75 | PUH | 0.814(0.791-0.836) | **0.879(0.861-0.897)** | 0.317(0.281-0.355) | **0.513(0.470-0.562)** |
| | UHB | 0.822(0.801-0.845) | **0.850(0.832-0.867)** | 0.271(0.243-0.302) | **0.324(0.290-0.367)** |
| | BH | 0.869(0.834-0.898) | **0.936(0.915-0.954)** | 0.552(0.483-0.623) | **0.710(0.643-0.785)** |
| | OUH "wave 2" | 0.822(0.811-0.832) | **0.867(0.858-0.875)** | 0.440(0.422-0.458) | **0.603(0.584-0.622)** |
| | OUH "wave 1" | 0.802(0.784-0.819) | **0.837(0.822-0.853)** | 0.063(0.056-0.070) | **0.095(0.082-0.112)** |

Supplementary Table 6: Comparison of AUROC and AUPRC performances using a XGBoost model (alongside 95% CIs), between same label and mixed label patient sample combinations, for the CURIAL datasets.

| N | Test Set | AUROC | | AUPRC | |
|---|---|---|---|---|---|
| | | Same Labels | Mixed Labels | Same Labels | Mixed Labels |
| Baseline (N=1) | PUH | 0.892(0.873-0.908) | | 0.606(0.565-0.647) | |
| | UHB | 0.841(0.823-0.859) | | 0.33(0.294-0.373) | |
| | BH | 0.914(0.89-0.933) | | 0.699(0.638-0.763) | |
| | OUH "wave 2" | 0.856(0.847-0.865) | | 0.597(0.578-0.616) | |
| | OUH "wave 1" | 0.818(0.802-0.834) | | 0.152(0.13-0.179) | |
| 5 | PUH | **0.897(0.881-0.912)** | 0.894(0.875-0.91) | 0.543(0.501-0.588) | **0.583(0.54-0.631)** |
| | UHB | 0.867(0.851-0.882) | 0.867(0.851-0.882) | **0.39(0.353-0.432)** | 0.374(0.335-0.418) |
| | BH | **0.925(0.906-0.944)** | 0.918(0.899-0.937) | **0.71(0.646-0.777)** | 0.673(0.606-0.737) |
| | OUH "wave 2" | **0.875(0.867-0.883)** | 0.869(0.86-0.877) | **0.611(0.592-0.629)** | 0.608(0.589-0.626) |
| | OUH "wave 1" | **0.873(0.861-0.886)** | 0.837(0.823-0.851) | **0.172(0.148-0.199)** | 0.148(0.126-0.176) |
| 25 | PUH | 0.872(0.854-0.888) | **0.882(0.864-0.901)** | 0.404(0.362-0.45) | **0.574(0.527-0.619)** |
| | UHB | 0.843(0.826-0.859) | **0.858(0.842-0.874)** | 0.289(0.254-0.33) | **0.312(0.275-0.353)** |
| | BH | 0.871(0.84-0.899) | **0.892(0.866-0.919)** | 0.606(0.537-0.677) | **0.647(0.575-0.713)** |
| | OUH "wave 2" | 0.848(0.839-0.856) | **0.859(0.85-0.867)** | 0.495(0.475-0.515) | **0.551(0.532-0.571)** |
| | OUH "wave 1" | **0.864(0.851-0.876)** | 0.851(0.837-0.864) | 0.099(0.086-0.114) | **0.122(0.104-0.145)** |
| 50 | PUH | 0.861(0.844-0.877) | **0.886(0.868-0.904)** | 0.352(0.311-0.398) | **0.59(0.548-0.633)** |
| | UHB | 0.833(0.817-0.849) | **0.855(0.838-0.87)** | 0.258(0.226-0.297) | **0.324(0.286-0.364)** |
| | BH | 0.868(0.841-0.893) | **0.893(0.868-0.917)** | 0.54(0.47-0.615) | **0.651(0.582-0.719)** |
| | OUH "wave 2" | 0.827(0.818-0.834) | **0.858(0.849-0.866)** | 0.429(0.409-0.448) | **0.57(0.552-0.589)** |
| | OUH "wave 1" | 0.853(0.841-0.867) | **0.852(0.838-0.866)** | 0.08(0.07-0.094) | **0.134(0.115-0.158)** |
| 75 | PUH | 0.847(0.83-0.864) | **0.888(0.87-0.905)** | 0.331(0.292-0.375) | **0.581(0.539-0.623)** |
| | UHB | 0.823(0.805-0.839) | **0.853(0.836-0.868)** | 0.254(0.221-0.292) | **0.306(0.268-0.346)** |
| | BH | 0.844(0.816-0.871) | **0.891(0.864-0.916)** | 0.491(0.423-0.563) | **0.637(0.565-0.704)** |
| | OUH "wave 2" | 0.816(0.806-0.824) | **0.853(0.844-0.861)** | 0.418(0.399-0.437) | **0.547(0.528-0.566)** |
| | OUH "wave 1" | 0.838(0.824-0.853) | **0.84(0.826-0.854)** | 0.076(0.065-0.089) | **0.119(0.102-0.142)** |

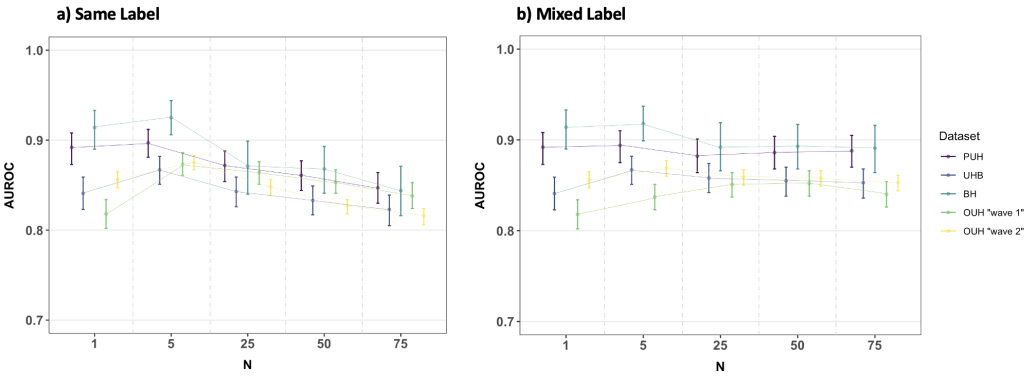


Supplementary Figure 1: AUROC performances using an XGBoost model, alongside 95% confidence intervals, for different values of N for (a) class-wise *GATS* and (b) mixed-class *GATS*. N=1 represents baseline performance on the original dataset. Results presented use a 3:1 ratio of same-label and mixed label *GATS*, respectively. Full numerical results for both AUROC and AUPRC can be found in the Supplementary Material.

Supplementary Table 7: Ablation of the mixed label ratio (i.e. the proportion of COVID-19 positive vs negative patients used to create mixed label *GATS* samples.

| pos:neg labels | Dataset | N=5 | N=50 | N=100 |
|---|---|---|---|---|
| 6:4 | PUH | 0.89(0.872-0.907) | 0.894(0.878-0.91) | 0.887(0.869-0.905) |
| | UHB | 0.863(0.844-0.881) | 0.865(0.848-0.883) | 0.863(0.846-0.881) |
| | BH | 0.932(0.909-0.951) | 0.928(0.906-0.947) | 0.931(0.91-0.95) |
| | OUH "wave 2" | 0.873(0.864-0.881) | 0.87(0.861-0.878) | 0.877(0.869-0.885) |
| | OUH "wave 1" | 0.851(0.837-0.865) | 0.847(0.833-0.862) | 0.845(0.83-0.86) |
| 7:3 | PUH | 0.897(0.881-0.912) | 0.888(0.869-0.905) | 0.886(0.869-0.903) |
| | UHB | 0.871(0.853-0.886) | 0.865(0.847-0.882) | 0.851(0.833-0.869) |
| | BH | 0.939(0.919-0.955) | 0.91(0.882-0.933) | 0.931(0.91-0.95) |
| | OUH "wave 2" | 0.881(0.873-0.889) | 0.876(0.868-0.884) | 0.871(0.862-0.88) |
| | OUH "wave 1" | 0.86(0.847-0.873) | 0.851(0.837-0.866) | 0.85(0.836-0.864) |
| 8:2 | PUH | 0.896(0.878-0.912) | 0.887(0.869-0.904) | 0.882(0.864-0.899) |
| | UHB | 0.865(0.847-0.882) | 0.868(0.851-0.884) | 0.849(0.83-0.867) |
| | BH | 0.929(0.909-0.947) | 0.926(0.904-0.946) | 0.921(0.897-0.943) |
| | OUH "wave 2" | 0.872(0.863-0.881) | 0.875(0.867-0.884) | 0.866(0.857-0.875) |
| | OUH "wave 1" | 0.846(0.832-0.861) | 0.847(0.833-0.861) | 0.829(0.813-0.846) |
| 9:1 | PUH | 0.89(0.872-0.907) | 0.874(0.855-0.893) | 0.87(0.85-0.889) |
| | UHB | 0.859(0.84-0.876) | 0.842(0.822-0.862) | 0.856(0.839-0.873) |
| | BH | 0.931(0.91-0.951) | 0.926(0.902-0.947) | 0.922(0.9-0.943) |
| | OUH "wave 2" | 0.872(0.863-0.88) | 0.865(0.856-0.874) | 0.866(0.857-0.875) |
| | OUH "wave 1" | 0.846(0.831-0.861) | 0.823(0.806-0.84) | 0.843(0.829-0.857) |

Supplementary Table 8: Ablation of the batch size used to generate *GATS* samples.

| batch size | Dataset | N=5 | N=50 | N=100 |
|---|---|---|---|---|
| 128 | PUH | 0.897(0.881-0.913) | 0.881(0.861-0.899) | 0.879(0.86-0.897) |
| | UHB | 0.87(0.854-0.886) | 0.854(0.836-0.872) | 0.854(0.835-0.873) |
| | BH | 0.928(0.908-0.945) | 0.918(0.889-0.942) | 0.928(0.907-0.947) |
| | OUH "wave 2" | 0.872(0.863-0.88) | 0.878(0.87-0.886) | 0.873(0.865-0.881) |
| | OUH "wave 1" | 0.845(0.83-0.859) | 0.846(0.831-0.862) | 0.851(0.837-0.866) |
| 256 | PUH | 0.896(0.88-0.912) | 0.887(0.869-0.904) | 0.888(0.871-0.906) |
| | UHB | 0.865(0.848-0.881) | 0.857(0.839-0.875) | 0.864(0.847-0.88) |
| | BH | 0.932(0.912-0.949) | 0.912(0.885-0.935) | 0.912(0.886-0.936) |
| | OUH "wave 2" | 0.875(0.866-0.883) | 0.872(0.863-0.881) | 0.875(0.866-0.883) |
| | OUH "wave 1" | 0.863(0.85-0.876) | 0.846(0.832-0.86) | 0.853(0.84-0.867) |
| 512 | PUH | 0.888(0.869-0.905) | 0.882(0.863-0.9) | 0.886(0.869-0.904) |
| | UHB | 0.865(0.847-0.882) | 0.848(0.829-0.866) | 0.861(0.842-0.879) |
| | BH | 0.931(0.911-0.95) | 0.922(0.897-0.943) | 0.929(0.906-0.947) |
| | OUH "wave 2" | 0.879(0.871-0.887) | 0.868(0.859-0.876) | 0.871(0.862-0.879) |
| | OUH "wave 1" | 0.852(0.838-0.867) | 0.851(0.837-0.866) | 0.854(0.84-0.868) |
| 1024 | PUH | 0.89(0.872-0.908) | 0.891(0.875-0.907) | 0.894(0.876-0.91) |
| | UHB | 0.862(0.844-0.88) | 0.858(0.84-0.875) | 0.874(0.857-0.891) |
| | BH | 0.931(0.91-0.95) | 0.928(0.905-0.947) | 0.936(0.917-0.954) |
| | OUH "wave 2" | 0.878(0.869-0.886) | 0.868(0.86-0.877) | 0.887(0.878-0.894) |
| | OUH "wave 1" | 0.853(0.839-0.867) | 0.848(0.832-0.862) | 0.861(0.847-0.876) |
| 2048 | PUH | 0.897(0.881-0.912) | 0.888(0.869-0.905) | 0.886(0.869-0.903) |
| | UHB | 0.871(0.853-0.886) | 0.865(0.847-0.882) | 0.851(0.833-0.869) |
| | BH | 0.939(0.919-0.955) | 0.91(0.882-0.933) | 0.931(0.91-0.95) |
| | OUH "wave 2" | 0.881(0.873-0.889) | 0.876(0.868-0.884) | 0.871(0.862-0.88) |
| | OUH "wave 1" | 0.86(0.847-0.873) | 0.851(0.837-0.866) | 0.85(0.836-0.864) |

Supplementary Table 9: Ablation of the proportion of mixed label *GATS* samples added to training.

| p | Dataset | N=5 | N=50 | N=100 |
|---|---|---|---|---|
| 0.9 | PUH | 0.891(0.874-0.908) | 0.897(0.882-0.912) | 0.887(0.87-0.904) |
| | UHB | 0.854(0.835-0.874) | 0.859(0.841-0.876) | 0.856(0.837-0.873) |
| | BH | 0.934(0.913-0.952) | 0.929(0.908-0.948) | 0.929(0.91-0.946) |
| | OUH "wave 2" | 0.875(0.866-0.883) | 0.874(0.865-0.882) | 0.876(0.868-0.884) |
| | OUH "wave 1" | 0.843(0.829-0.859) | 0.855(0.842-0.868) | 0.848(0.834-0.863) |
| 0.7 | PUH | 0.901(0.885-0.916) | 0.892(0.874-0.909) | 0.887(0.868-0.904) |
| | UHB | 0.864(0.846-0.881) | 0.865(0.848-0.881) | 0.855(0.838-0.874) |
| | BH | 0.93(0.91-0.95) | 0.928(0.907-0.949) | 0.915(0.888-0.938) |
| | OUH "wave 2" | 0.878(0.869-0.886) | 0.876(0.868-0.884) | 0.87(0.862-0.878) |
| | OUH "wave 1" | 0.853(0.84-0.867) | 0.856(0.843-0.871) | 0.847(0.832-0.861) |
| 0.5 | PUH | 0.892(0.875-0.909) | 0.886(0.869-0.904) | 0.889(0.872-0.906) |
| | UHB | 0.861(0.842-0.879) | 0.862(0.846-0.879) | 0.863(0.845-0.881) |
| | BH | 0.935(0.915-0.953) | 0.933(0.913-0.95) | 0.923(0.901-0.942) |
| | OUH "wave 2" | 0.878(0.87-0.886) | 0.874(0.866-0.882) | 0.875(0.867-0.883) |
| | OUH "wave 1" | 0.849(0.834-0.862) | 0.852(0.837-0.866) | 0.85(0.837-0.865) |
| 0.3 | PUH | 0.89(0.873-0.907) | 0.89(0.873-0.907) | 0.888(0.871-0.905) |
| | UHB | 0.861(0.843-0.878) | 0.859(0.841-0.876) | 0.861(0.844-0.879) |
| | BH | 0.937(0.918-0.953) | 0.93(0.909-0.95) | 0.919(0.895-0.939) |
| | OUH "wave 2" | 0.877(0.868-0.885) | 0.879(0.871-0.887) | 0.868(0.86-0.876) |
| | OUH "wave 1" | 0.853(0.84-0.867) | 0.861(0.848-0.874) | 0.845(0.831-0.86) |
| 0.1 | PUH | 0.892(0.875-0.908) | 0.878(0.86-0.896) | 0.885(0.867-0.903) |
| | UHB | 0.867(0.849-0.884) | 0.862(0.845-0.878) | 0.856(0.839-0.872) |
| | BH | 0.931(0.91-0.95) | 0.924(0.901-0.943) | 0.918(0.893-0.94) |
| | OUH "wave 2" | 0.868(0.859-0.876) | 0.875(0.866-0.883) | 0.872(0.864-0.88) |
| | OUH "wave 1" | 0.845(0.83-0.86) | 0.843(0.83-0.857) | 0.84(0.825-0.855) |

##### D.2 eICU dataset

Supplementary Table 10: Comparison of AUROC and AUPRC performances using a neural network (alongside 95% CIs), for the eICU dataset.

| N | AUROC | | AUPRC | |
|---|---|---|---|---|
| | Same Labels | Mixed Labels | Same Labels | Mixed Labels |
| Baseline (N=1) | 0.851(0.843-0.86) | | 0.473(0.453-0.495) | |
| 5 | **0.857(0.849-0.865)** | 0.856(0.848-0.864) | **0.468(0.448-0.491)** | 0.463(0.442-0.486) |
| 25 | 0.837(0.829-0.845) | **0.848(0.84-0.856)** | 0.344(0.328-0.361) | **0.455(0.435-0.478)** |
| 50 | 0.822(0.814-0.831) | **0.847(0.839-0.855)** | 0.319(0.303-0.335) | **0.448(0.428-0.47)** |
| 75 | 0.802(0.792-0.811) | **0.846(0.838-0.855)** | 0.291(0.276-0.306) | **0.449(0.429-0.471)** |
| 100 | 0.781(0.771-0.791) | **0.849(0.841-0.857)** | 0.27(0.256-0.284) | **0.454(0.434-0.476)** |

Supplementary Table 11: Comparison of AUROC and AUPRC performances using XGBoost (alongside 95% CIs), for the eICU dataset.

| N | AUROC | | AUPRC | |
|---|---|---|---|---|
| | Same Labels | Mixed Labels | Same Labels | Mixed Labels |
| Baseline (N=1) | 0.879(0.872-0.886) | | 0.527(0.507-0.549) | |
| 5 | 0.809(0.8-0.819) | **0.864(0.857-0.872)** | 0.367(0.347-0.387) | **0.495(0.475-0.517)** |
| 25 | 0.762(0.751-0.772) | **0.864(0.857-0.872)** | 0.272(0.256-0.291) | **0.494(0.474-0.516)** |
| 50 | 0.751(0.74-0.761) | **0.864(0.856-0.872)** | 0.248(0.231-0.265) | **0.499(0.478-0.52)** |
| 75 | 0.740(0.73-0.751) | **0.864(0.857-0.872)** | 0.237(0.222-0.253) | **0.496(0.475-0.519)** |
| 100 | 0.741(0.731-0.752) | **0.867(0.86-0.875)** | 0.235(0.22-0.251) | **0.501(0.482-0.522)** |

##### D.3 Adult (Census Income) dataset

Supplementary Table 12: Comparison of AUROC and AUPRC performances using a neural network model (alongside 95% CIs), for the Adult dataset.

| N | AUROC | | AUPRC | |
|---|---|---|---|---|
| | Same Labels | Mixed Labels | Same Labels | Mixed Labels |
| Baseline (N=1) | 0.888(0.882-0.895) | | 0.736(0.722-0.753) | |
| 5 | 0.858(0.851-0.866) | **0.877(0.870-0.884)** | 0.664(0.647-0.682) | **0.685(0.666-0.704)** |
| 25 | 0.838(0.831-0.846) | **0.869(0.862-0.876)** | 0.607(0.589-0.627) | **0.704(0.687-0.721)** |
| 50 | 0.827(0.82-0.835) | **0.873(0.866-0.880)** | 0.538(0.522-0.558) | **0.692(0.674-0.710)** |
| 75 | 0.823(0.816-0.831) | **0.878(0.871-0.886)** | 0.506(0.49-0.524) | **0.709(0.692-0.727)** |
| 100 | 0.819(0.812-0.827) | **0.871(0.864-0.878)** | 0.497(0.482-0.514) | **0.688(0.669-0.707)** |

Supplementary Table 13: Comparison of AUROC and AUPRC performances using a XGBoost model (alongside 95% CIs), for the Adult dataset.

| N | AUROC | | AUPRC | |
|---|---|---|---|---|
| | Same Labels | Mixed Labels | Same Labels | Mixed Labels |
| Baseline (N=1) | 0.928(0.922-0.933) | | 0.826(0.814-0.839) | |
| 5 | 0.883(0.877-0.889) | **0.909(0.904-0.915)** | 0.689(0.671-0.709) | **0.789(0.774-0.802)** |
| 25 | 0.83(0.822-0.839) | **0.911(0.905-0.917)** | 0.629(0.610-0.649) | **0.790(0.776-0.804)** |
| 50 | 0.783(0.774-0.793) | **0.913(0.907-0.919)** | 0.534(0.516-0.556) | **0.797(0.784-0.811)** |
| 75 | 0.739(0.728-0.749) | **0.913(0.907-0.919)** | 0.448(0.431-0.468) | **0.796(0.783-0.81)** |
| 100 | 0.736(0.725-0.746) | **0.913(0.908-0.919)** | 0.455(0.436-0.475) | **0.798(0.785-0.811)** |
